# Binge Eating and Psychological Distress: Investigating Relationships with Depression, Anxiety, and Stress

**DOI:** 10.1101/2025.01.24.25321068

**Authors:** Ana Carolina Soares Marinho, Julio Cezar Albuquerque da Costa, Sheyla C.S Fernandes

## Abstract

Binge eating (BE) involves the consumption of a greater amount of food than most people would under similar conditions, within a specified period and may be associated with various psychiatric disorders. This study aimed to compare the variables of Depression, Anxiety, and Stress among groups defined by the severity of binge eating symptoms. Participants were recruited through social media advertisements and responded online to the Depression, Anxiety, and Stress Scale (DASS-21) and the Binge Eating Scale (BES). It was observed that individuals with medium-high depressive symptoms showed a higher presence of binge eating symptoms, while those with moderate, severe, and extremely severe anxiety symptoms exhibited an increased occurrence of binge eating. The findings suggest a need for interventions addressing binge eating with comorbidities such as depression and anxiety, or binge eating as a comorbidity of these conditions.

## Introduction

### Eating Disorders

Eating behaviors reflect an individual’s relationship with food and their behavioral responses to food-related cues in the environment (Russell & Russel, 2018). These behaviors develop early and follow different trajectories (Herle et al., 2020). In children, it is common to observe tendencies such as overeating, undereating, eating only specific foods, and refusing to try new ones. Such behaviors may serve as precursors to dieting during adolescence, which is a risk factor for eating disorders (ED) (Herle et al., 2020; Thornton et al., 2017). Furthermore, adolescence and early adulthood are critical periods of vulnerability for the development of EDs (Klein, Sylvester & Schvey, 2021).

EDs are conditions characterized by a disturbed eating pattern and weight control behaviors that impact an individual’s physical health and social functioning (Treasure et al., 2020). Although their etiology remains unknown and likely involves biological, psychological, and environmental factors, stressful experiences are widely recognized as risk factors for the development and maintenance of EDs (Rojo et al., 2006). According to the 5th edition of the Diagnostic and Statistical Manual of Mental Disorders (DSM-5), EDs include: Anorexia Nervosa, Bulimia Nervosa, Binge-Eating Disorder (BED), Avoidant/Restrictive Food Intake Disorder, Rumination Disorder, and Pica (APA, 2022).

Anorexia nervosa is characterized by food restriction, leading to low body weight; an intense fear of gaining weight or becoming “fat”; and body image distortion. Bulimia nervosa, on the other hand, is characterized by binge eating (consumption of an unusually large amount of food within a two-hour period, accompanied by a sense of loss of control); unhealthy weight control behaviors such as vomiting, laxative use, dietary restriction, or excessive exercise; and self-evaluation being highly influenced by body shape. Additionally, binge-eating disorder is characterized by recurrent binge-eating episodes without the use of compensatory weight-control behaviors (Klein, Sylvester & Schvey, 2021).

### Binge-Eating Disorder

Binge-Eating Disorder (BED) is characterized by recurrent binge-eating episodes with fewer compensatory behaviors compared to Bulimia Nervosa (Treasure et al., 2020). BED is associated with various psychological and non-psychological difficulties that impair daily life functioning (Iqbal & Rehman, 2019). It results from a combination of psychological, social, cultural, and biological factors, with potential risk factors including childhood obesity, perfectionism, conduct problems, substance abuse, body image distortion, and involvement of dopamine and μ-opioid receptors, among others (Hilbert, 2018). Moreover, Herle et al. (2020) observed that childhood overeating is associated with an increased risk of developing BED by the age of 16.

Emotional, social, and cognitive dysfunctions have been identified as maintenance factors for BED (Hilbert, 2018), with binge-eating episodes often occurring in a context of reduced emotional awareness and difficulties in emotional regulation (Dingemans, Danner & Parks, 2017). These findings align with the study by Fernandes, Marinho & Costa (2023), which found that individuals who experience binge-eating episodes perceive factors such as “emotional dysregulation,” “lack of self-control,” and “low emotional intelligence” as facilitators of binge eating.

### Binge Eating and Psychological Distress

The COVID-19 pandemic exposed individuals to various stressors, such as social isolation, economic difficulties, routine disruption, and fear of infection (Wasserman et al., 2020). Reactions to stress, including attention difficulties, insomnia, irritability, and interpersonal conflicts, increased during the pandemic (Brooks et al., 2020). There was also a rise in pathological anxiety and depression, particularly in populations such as the Chinese (Li et al., 2020). Additionally, individuals with EDs exhibited increased food restriction, physical exercise, binge-eating, and purging episodes, as well as worsening of both specific ED symptoms and internalizing symptoms (such as anxiety, depression, and post-traumatic stress symptoms) (Castellini et al., 2020; Monteleone et al., 2021).

The escape theory, proposed by Heatherton & Baumeister (1991), suggests that binge eating provides an “escape,” temporarily allowing individuals to dissociate from negative affective experiences. According to Walenda et al. (2021), “negative affect” includes several components, such as depressed mood, sadness, anxiety, and anger. This theory helps illustrate the relationship between binge eating and conditions such as anxiety, depression, and stress. For example, studies suggest that binge eating serves as a coping mechanism for anxiety (Binford et al., 2004; Mitchell et al., 1999). However, while binge eating may offer temporary relief from negative affect, it can lead to increased levels of depression and shame, further increasing the likelihood of another binge episode (Stice, 1998). Furthermore, according to Freeman & Gil (2004), high psychological stress is associated with binge-eating episodes, and elevated stress levels can interact with anxiety, increasing the risk of binge-eating tendencies (Jung et al., 2017). BED is also comorbid with several psychiatric disorders, the most common being mood and anxiety disorders. In a study by Grilo, White, and Masheb (2009) involving 404 patients with BED, 73.8% had a history of psychiatric disorders, and 43.1% had at least one current psychiatric disorder. In this study, the most common disorders in the patients’ history were mood, anxiety, and substance use disorders, while the most prevalent current disorders were mood and anxiety disorders. Thus, the present study aimed to compare the variables of depression, anxiety, and stress among groups categorized based on the severity of binge-eating symptoms.

## Methods

### Design and Participants

The research was conducted through the application of the selected instruments. A total of 80 research participants met the inclusion criteria: 1) Over 18 years old; 2) Literate; and 3) Having experienced at least three binge eating episodes in the past year. The sample size was calculated using the G*Power software (Faul et al., 2009). Participants were invited to take part in the study through social media posts on Instagram, Facebook, WhatsApp, and Twitter. It was explained that binge eating is characterized by “eating an amount of food that is definitely larger than most people would eat in a similar period under similar circumstances” and “a sense of lack of control over eating” (APA, 2014).

Of the 80 research participants, the majority self-identified as female (n=64, 80.00%), White (n=55, 68.75%), with a high school diploma (n=38, 60.00%), and a household income of 1 to 3 minimum wages (n=21, 26.25%), with ages ranging from 19 to 45 years (M=25.24, SD=5.61). The demographic profile of the sample is presented in Table 1.

### Instruments

1. Depression, Anxiety, and Stress Scale (DASS-21) – This instrument was developed by Lovibond (1995) and adapted to the Brazilian context by Vignola and Tucci (2014). It consists of 21 items divided into three subscales that assess the intensity of behaviors and feelings experienced over the past week: 1) Depression (e.g., “I couldn’t experience any positive feelings”); 2) Stress (e.g., “I found it hard to calm down”); and 3) Anxiety (e.g., “I worried about situations where I might panic and appear foolish”). The instrument follows a 4-point Likert scale, ranging from 0 (does not apply to me) to 3 (applies very much to me). It has good internal consistency for each scale, with Cronbach’s alpha of .92 for depression, .90 for stress, and .86 for anxiety (Vignola & Tucci, 2014).
2. Binge Eating Scale (BES) – Originally developed by Gormally et al. (1982) and adapted to the Brazilian context by Freitas et al. (2001). This Likert-scale instrument consists of 16 items and 62 statements assessing the severity of binge eating disorder based on: 1) Binge eating characteristics (16 items, including 8 behavioral manifestations and 8 emotional and cognitive aspects); 2) Statements indicating the severity of each characteristic (ranging from 0 to 3 points); and 3) Three dimensions to assess binge eating severity (frequency, amount of food consumed, and emotional involvement in an episode) (Freitas et al., 2001). The instrument has a moderately high internal consistency, with Cronbach’s alpha of .85 (Gormally et al., 1982).

### Procedures

Research participants were recruited through social media advertisements. Upon choosing to participate in the study, they only needed to click on the provided link, which directed them to the Google Forms platform. Data collection took place on Google Forms, making the research instruments available entirely online. Additionally, the platform included the Informed Consent Form (ICF) and the Sociodemographic Questionnaire.

This study was submitted to and approved by the Ethics Committee of the Federal University of Alagoas under opinion number 5.631.322 and CAAE 60175922.5.0000.5013. All participants accessed the Informed Consent Form and voluntarily agreed to participate in the study.

### Data Analysis

For data processing and quantitative analysis, Excel software was used for data tabulation, and JASP 0.17.1 software (JASP, 2022) was employed for statistical analysis. To test for data normality, the Shapiro-Wilk test was applied, where statistically significant values (p > 0.05) reject the null hypothesis of data normality, indicating a skewed distribution.

Based on this, the Kruskal-Wallis test (1952) was used to compare binge eating levels (Dependent Variable) according to participants’ levels of depression, anxiety, and stress (Independent Variables). The cutoff points followed the recommendations of Lovibond & Lovibond (2014), categorizing participants into normal, mild, moderate, severe, and extremely severe symptom groups. Furthermore, Dunn’s test (Dunn, 1964) was conducted for pairwise group comparisons as a post-hoc analysis.

Finally, to investigate the influence of psychological distress indicators on binge eating, multiple linear regression analysis was performed, considering Depression, Anxiety, and Stress as predictor variables and binge eating as the outcome variable. The Stepwise method was employed to formulate statistical models with predictor variables that significantly influenced the outcome variable, excluding those without statistical significance (Field, 2013).

## Results

Initially, the normality analysis of the data was conducted to investigate whether the sample resulted in a symmetrical distribution. The results indicated a non-normal distribution. Furthermore, the results of the Shapiro-Wilk test, which is recommended for samples with more than 50 observations (Field, 2005), confirmed the asymmetry of the data (Shapiro-Wilk (80) = 0.955, p = 0.007). Therefore, the inferential statistical tests used in this study were non-parametric.

Additionally, the sample scores on the studied constructs were analyzed using descriptive statistics (mean, standard deviation, and median), along with the cutoff points adopted by the instruments (Vignola & Tucci, 2014). The results showed that the overall sample obtained mild depression scores, moderate anxiety scores, normal stress levels, and moderate binge eating scores.

The levels of the DASS-21 were also investigated as independent groups to assess the equivalence of observations across the groups used in the descriptive statistics.

Regarding the depression variable, 45% (n = 36) of participants presented normal symptoms, 10% (n = 8) had mild symptoms, 35% (n = 28) presented moderate symptoms, and 10% (n = 8) exhibited severe symptoms. No participants were classified as having extremely severe symptoms. Thus, four groups were defined for comparative analysis.

For the anxiety variable, 23.75% (n = 19) of participants had normal symptoms, 10% (n = 8) had mild symptoms, 27.5% (n = 22) presented moderate symptoms, 26.25% (n = 21) had severe symptoms, and 12.50% (n = 10) exhibited extremely severe symptoms.

Regarding stress levels, low variability was observed, with most observations (81.25%, n = 65) falling within the normal level, contrasting with the mild (11.25%, n = 9) and moderate (7.50%, n = 6) levels. No participants scored in the severe or extremely severe stress categories.

Binge Eating Disorder (BED) severity was assessed using the BES instrument scores. In this regard, 40% (n = 32) of the individuals did not exhibit BED, while 27.5% (n = 22) had moderate BED, and 32.5% (n = 26) had severe BED. It is important to highlight that the comparative analysis used the overall instrument score, calculated by summing the participant’s responses, which were recalculated based on the variables proposed by Freitas et al. (2001).

### Group Comparison

The results of the comparative tests indicated statistically significant differences among participants with depression symptoms [H(3) = 13.104, p = 0.004, η^2^ = 0.165], as shown in Figure 1. Additionally, Dunn’s test indicated differences between levels 1 (normal) and 3 (moderate) of depression (z = -2.905, p = 0.004), as well as between levels 1 (normal) and 4 (severe) (z = -2.740, p = 0.06). This suggests that individuals with normal depression levels scored statistically differently compared to those with moderate and severe levels of the disorder. Therefore, it can be inferred that the presence of moderate to high depressive symptoms is associated with more frequent eating disorder symptoms.

The analysis of anxiety symptoms also revealed statistically significant differences based on the Kruskal-Wallis test [H(4) = 16.8, p = 0.002, η^2^ = 0.212]. It was found that level 1 (normal) differed significantly from level 4 (severe) (z = -2.505, p = 0.012) and level 5 (extremely severe) (z = -3.137, p = 0.002). Furthermore, level 2 (mild) showed significant differences compared to levels 4 (severe) (z = -2.625, p = 0.009) and 5 (extremely severe) (z = -3.210, p = 0.001). Descriptive analysis indicates a decrease in binge eating for normal and mild anxiety groups, but an increase in BED scores in the moderate, severe, and extremely severe anxiety levels.

The results for stress symptom groups indicated no statistically significant differences between the groups [H(2) = 4.627, p = 0.099, η^2^ = 0.054]. However, this result may be related to the measurement residual, where the confidence interval for level 3 was much larger than for the other levels. Moreover, the sample exhibited less variability in stress levels compared to depression and anxiety.

Finally, the influence of psychological distress indicators on Binge Eating Disorder was tested using a multiple linear regression analysis, adopting the Stepwise method. The best-fitting model [F(2;77)= 11.989, p < 0.001, adjusted R^2^ = 0.218] included the variables Depression (β = 0.302, t = 2.603, p = 0.011) and Anxiety (β = 0.257, t = 2.212, p = 0.030), explaining 21.8% of the data variability. Thus, the stress variable did not significantly influence BED (β = 0.020, t = 0.177, p = 0.860).

Based on this, it can be inferred that an increase of 1 standard deviation (SD) in the Depression variable resulted in an increase of 0.302 SD in BED, while an increase of 1 SD in Anxiety directly impacted a 0.257 SD increase in BED. In contrast, stress levels did not directly impact BED severity.

## Discussion

The analysis results suggest a trend in which individuals with moderate to high depressive symptoms tend to exhibit a greater presence of binge eating (BE) symptoms. In contrast, individuals with normal and mild anxiety show a decrease in BE, while those with moderate, severe, and extremely severe anxiety symptoms show an increase in BE. This finding aligns with other studies, such as Albuquerque, Bahia, and Maynard (2021), which demonstrate a statistically significant relationship between anxiety, depression, and binge eating. Additionally, the study by Grilo, White, and Masheb (2009) highlighted the prevalence of mood and anxiety disorders in patients with eating disorders (ED).

Furthermore, although various psychiatric comorbidities are associated with ED, mood and anxiety disorders appear at higher levels. In this context, the associations between anxiety, depression, and binge eating found in our study may be related to common risk factors and etiology (Garcia et al., 2020).

However, the findings indicate no significant relationship between BE symptoms and stress symptoms, which contrasts with findings from studies such as Pike et al. (2006), which reported that ED patients experienced a higher number of stressful situations compared to the control group. On the other hand, in the study by Smith et al. (2021), BE symptoms were associated with accumulated stress (accumulation of stressors and responses) but not with reactive stress (related to the magnitude of the initial response) and recovery stress (related to the time it takes for stress levels to return to baseline). Thus, assessing the “stress” factor more specifically may be related to different results. Additionally, our findings contrast with statements from the APA (2022), which cites “interpersonal stressors” as a potential trigger for BE, including being teased or victimized by peers, difficulties with communication and affiliation, and mistrust of others (Schell & Racine, 2023). However, once again, this would be a specific aspect of stress that was not evaluated in our study, as the DASS-21 stress scale refers to aspects such as difficulty relaxing, tension, irritability, and agitation (Lovibond & Lovibond, 1995).

Moreover, factors such as education and income may also interfere with the results of this study, as they are related to the development of common mental disorders (signs and symptoms of insomnia, fatigue, irritability, forgetfulness, difficulty concentrating, and somatic complaints) (Goldberg & Huxley, 1992, as cited in Ludermir & Filho, 2002), potentially affecting the constructs of depression, anxiety, and stress. In the study by Ludermir & Filho (2002), it was observed that individuals with up to four years of education and those with a per capita family income of up to ¼ of the minimum wage were 2.84 and 2.87 times more likely, respectively, to present common mental disorders than those with 11 or more years of education and with a per capita family income greater than one minimum wage. However, the population in our study consisted entirely of individuals with more than 11 years of education, and the majority had a family income between 1 and 3 minimum wages, thus representing a sample with little variability and representativeness. Additionally, the factor of race also proves to be relevant when associated with common mental disorders. The study by Anselmi et al. (2008) observed a higher prevalence of these symptoms in Black individuals, especially Black women, similar to the study by Campos et al. (2020), which pointed to these signs as more prevalent in Black women, followed by non-Black women, Black men, and non-Black men. However, in our study, only 5% of respondents were Black individuals.

Nonetheless, it is believed that the presented results are justified by the sample nature of this study, which did not include a clinically diagnosed sample, as well as the reduced sample size, showing little variability. However, the study by Albuquerque, Bahia, and Maynard (2021) also did not include a clinically diagnosed population and had a similarly small sample size, with 93 participants. Thus, although it presents similar results to our study, it also has similar limitations, despite using the BES instrument. In their study, 74.2% of participants fell into the “No BE” category, whereas in our study, only 40% of participants fell into this category. On the other hand, the study by Pike et al. (2006) included a clinically diagnosed ED population with a sample size of 162 ED participants and 162 control group participants, as well as another group with 107 individuals with another psychiatric diagnosis. Additionally, the study by Grilo, White, and Masheb (2009) also included a clinically diagnosed ED population with a larger sample size of 404 volunteer participants. Thus, it is considered that the studies by Pike et al. (2006) and Grilo, White, and Masheb (2009) present greater sample power among those cited.

## Final Considerations

This study aimed to compare the variables of depression, anxiety, and stress among groups defined by the severity of psychological distress symptoms. The findings suggest a potential need for interventions that consider binge eating (BE) alongside comorbidities such as depression and anxiety, or BE as a comorbidity of these conditions.

However, the present study has limitations. First, the convenience sampling method used in data collection poses a constant challenge regarding sample representativeness (Marconi & Lakatos, 2017). Additionally, it is recommended that future studies replicate the research using systematic samples and a longitudinal approach, incorporating control groups such as individuals undergoing psychotherapy, those using psychotropic medications, and other clinical populations. This would allow for a more in-depth investigation into the effectiveness of interventions in addressing psychological distress and BE. Furthermore, conducting similar studies in different regions of Brazil with greater exploration of variables such as income, gender, and race is essential to provide a broader understanding of the phenomenon.

## Data Availability

The complete dataset supporting the results of this study is available upon request from the corresponding author, Ana Carolina Marinho. The dataset is not publicly available due to privacy concerns related to the participants’ information.

## APPENDIX A

**Table 1.** Sociodemographic profile of the sample.

| Variables |  | n | % |
| --- | --- | --- | --- |
| Gender |  |  |  |
|  | Female | 64 | 80,00 |
|  | Male | 14 | 17,50 |
|  | Unidentified | 1 | 1,25 |
|  | Fluid | 1 | 1,25 |
| Race/Ethnicity |  |  |  |
|  | Yellow | 1 | 1,25 |
|  | White | 55 | 68,75 |
|  | Brown | 20 | 25,00 |
|  | Black | 4 | 5,00 |
| Education Level |  |  |  |
|  | High School Complete | 48 | 60,00 |
|  | College Complete | 24 | 30,00 |
|  | Postgraduate Complete | 8 | 10,00 |
| Family Income (BRL) |  |  |  |
|  | Up to 1 minimum wage | 4 | 5,00 |
|  | From 1 to 3 minimum wages | 21 | 26,25 |
|  | From 3 to 6 minimum wages | 19 | 23,75 |
|  | From 6 to 9 minimum wages | 14 | 17,50 |
|  | More than 9 minimum wages | 19 | 23,75 |
|  | No income | 3 | 3,75 |
| Disorder |  |  |  |
|  | Yes | 80 | 100,00 |
|  | No | 0 | 0,00 |

## APPENDIX B

**Figure 1.**
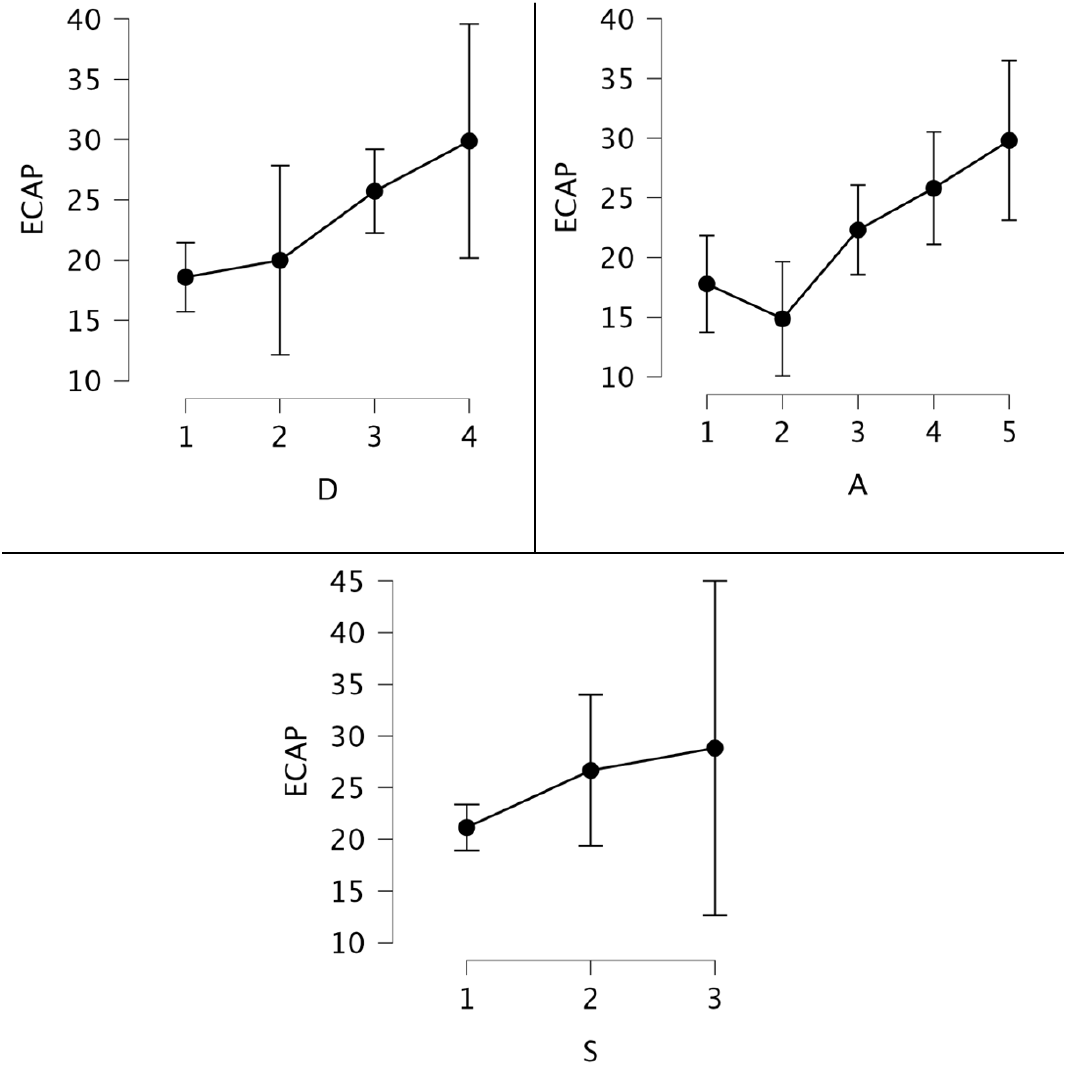
BES scores separated by levels of Depression, Anxiety, and Stress.

## References

Associação Americana de Psiquiatria. Manual Diagnóstico e Estatístico de Transtornos Mentais – DSM-5. 5^a^ ed. Porto Alegre: Artmed; 2014.

American Psychiatric Association. (2022). Diagnostic and statistical manual of mental disorders (5th ed., text rev.). 10.1176/appi.books.9780890425787

Albuquerque, A.L.,, Bahia, F.C.,, Maynard, D. Compulsão alimentar: uma análise da relação com os transtornos psicológicos da depressão e ansiedade. Research, Society and Development. Vol. 10. Num. 16. 2021. p.e380101623982–e380101623982

Anselmi, L., Barros, F. C., Minten, G. C., Gigante, D. P., Horta, B. L., & Victora, C. G. (2008). Prevalência e determinantes precoces dos transtornos mentais comuns na coorte de nascimentos de 1982, Pelotas, RS. Revista de Saúde Pública, 42, 26–33.

Binford, R. B., Pederson Mussell, M., Peterson, C. B., Crow, S. J., & Mitchell, J. E. (2004). Relation of binge eating age of onset to functional aspects of binge eating in binge eating disorder. International Journal of Eating Disorders, 35(3), 286–292.

Brooks, S.K.,, Webster, R.K.,, Smith, L.E.,, Woodland, L.,, Wessely, S.,, Greenberg, N.,, Rubin, G.J. The psychological impact of quarantine and how to reduce: Rapid review of the evidence. Lancet 2020, 395, 912–92

Campos, F. M., Araújo, T. M. D., Viola, D. N., Oliveira, P. C. S., & Sousa, C. C. D. (2020). Estresse ocupacional e saúde mental no trabalho em saúde: desigualdades de gênero e raça. Cadernos saude coletiva, 28, 579–589.

Castellini, G.,, Cassioli, E.,, Rossi, E.,, Innocenti, M.,, Gironi, V.,, Sanfilippo, G.,, Felciai, F.,, Monteleone, A.M.,, Ricca, V. The impact of COVID-19 epidemic on eating disorders: A longitudinal observation of pre versus post psychopathological features in a sample of patients with eating disorders and a group of healthy controls. Int. J. Eat. Disord. 2020, 53, 1855–1862.

Dingemans, A., Danner, U., & Parks, M. (2017). Emotion Regulation in Binge Eating Disorder: A Review. Nutrients, 9(11), 1274. 10.3390/nu9111274

Dunn, O. J. (1964). Multiple comparisons using rank sums. Technometrics, 6(3), 241–252.

Duchesne, M., Mattos, P., Appolinario, J.C., de Freitas, S.R., Coutinho, G., Santos, C., Coutinho, W., 2010. Assessment of executive functions in obese individuals with binge eating disorder. Rev. Bras. Psiquiatr. 32, 381–388.

Faul F, Erdfelder E, Buchner A, Lang AG. Statistical power analyses using G*Power 3.1: tests for correlation and regression analyses. Behav Res Methods. 2009; 41:1149–1160. 10.3758/BRM.41.4.1149.

Freeman, L. M. Y., & Gil, K. M. (2004). Daily stress, coping, and dietary restraint in binge eating. International Journal of Eating Disorders, 36(2), 204–212.

Garcia, S. C., Mikhail, M. E., Keel, P. K., Burt, S. A., Neale, M. C., Boker, S., & Klump, K. L. (2020). Increased rates of eating disorders and their symptoms in women with major depressive disorder and anxiety disorders. International Journal of Eating Disorders, 53(11), 1844–1854.

Grilo, C. M., White, M. A., & Masheb, R. M. (2009). DSM-IV psychiatric disorder comorbidity and its correlates in binge eating disorder. International Journal of Eating Disorders, 42(3), 228–234.

Goldberg, D. P., & Huxley, P. (1992). Common mental disorders: a bio-social model. Tavistock/Routledge.

Gormally, J. I. M., Black, S., Daston, S., & Rardin, D. (1982). The assessment of binge eating severity among obese persons. Addictive behaviors, 7(1), 47–55.

Hair, J. F., Black, W. C., Babin, B. J., Anderson, R. E., & Tatham, R. L. (2009). Análise multivariada de dados. Bookman editora.

Heatherton, T. F., & Baumeister, R. F. (1991). Binge eating as escape from self-awareness. Psychological bulletin, 110(1), 86.

Herle, M., Stavola, B. D., Hübel, C., Ferreira, D. L. S., Abdulkadir, M., Yilmaz, Z., … & Micali, N. (2020). Eating behavior trajectories in the first 10 years of life and their relationship with BMI. International journal of obesity, 44(8), 1766–1775.

Hilbert, Anja (2018). Binge-Eating Disorder. Psychiatric Clinics of North America, (), S0193953×18311699–. doi:10.1016/j.psc.2018.10.011

Hilbert, A., Pike, K. M., Wilfley, D. E., Fairburn, C. G., Dohm, F. A., & Striegel-Moore, R. H. (2011). Clarifying boundaries of binge eating disorder and psychiatric comorbidity: a latent structure analysis. Behaviour research and therapy, 49(3), 202–211.

Iqbal, A., & Rehman, A. (2019). Binge eating disorder

JASP Team (2023). JASP (Version 0.18.1)[Computer software]. https://jasp-stats.org/.

Jung, J. Y., Kim, K. H., Woo, H. Y., Shin, D. W., Shin, Y. C., Oh, K. S., … & Lim, S. W. (2017). Binge eating is associated with trait anxiety in Korean adolescent girls: a cross sectional study. BMC Women’s Health, 17(1), 1–7.

Kessler, R. C., Berglund, P. A., Chiu, W. T., Deitz, A. C., Hudson, J. I., Shahly, V., … & Xavier, M. (2013). The prevalence and correlates of binge eating disorder in the World Health Organization World Mental Health Surveys. Biological psychiatry, 73(9), 904–914.

Klein, D. A., Sylvester, J., & Schvey, N. A. (2021). Eating disorders in primary care: diagnosis and management. American family physician, 103(1), 22–32.

Kruskal, W. H., & Wallis, W. A. (1952). Use of ranks in one-criterion variance analysis. Journal of the American statistical Association, 47(260), 583–621.

Li, J.,, Yang, Z.,, Qiu, H.,, Wang, Y.,, Jian, L.,, Ji, J.,, Li, K. Anxiety and depression among general population in China at the peak of the COVID-19 epidemic. World Psychiatry 2020, 19, 249–250.

Lovibond, S. H. (1995). Manual for the depression anxiety stress scales. Sydney psychology foundation.

Lovibond, P. F., & Lovibond, S. H. (1995). The structure of negative emotional states: Comparison of the Depression Anxiety Stress Scales (DASS) with the Beck Depression and Anxiety Inventories. Behaviour research and therapy, 33(3), 335–343.

Lakatos, E. M., & Marconi, M. D. A. (2017). Metodologia do trabalho científico: projetos de pesquisa, pesquisa bibliográfica, teses de doutorado, dissertações de mestrado, trabalhos de conclusão de curso. São Paulo: Atlas.

Levitan, M. N., Papelbaum, M., Carta, M. G., Appolinario, J. C., & Nardi, A. E. (2021). Binge eating disorder: A 5-year retrospective study on experimental drugs. Journal of Experimental Pharmacology, 33–47.

Ludermir, A. B., & de Melo Filho, D.A. (2002). Condições de vida e estrutura ocupacional associadas a transtornos mentais comuns. Revista de Saúde Pública, 36, 213–221.

Mitchell JE, Mussell MP, Peterson CB, Crow S, Wonderlich SA, Crosby RD, Weller C. Hedonics of binge eating in women with bulimia nervosa and binge eating disorder. International Journal of Eating Disorders. 1999;26:165–170.

Monteleone, A.M.,, Marciello, F.,, Cascino, G.,, Abbate-Daga, G.,, Anselmetti, S.,, Baiano, M.,, Balestrieri, M.,, Barone, E.,, Bertelli, S.,, Carpiniello, B.,, et al. The impact of COVID-19 lockdown and of the following “re-opening” period on specific and general psychopathology in people with eating disorders: The emergent role of internalizing symptoms. J. Affect. Disord. 2021, 285, 77–83.

Pike, K. M., Wilfley, D., Hilbert, A., Fairburn, C. G., Dohm, F. A., & Striegel-Moore, R. H. (2006). Antecedent life events of binge-eating disorder. Psychiatry research, 142(1), 19–29.

Rojo, L.,, Conesa, L.,, Bermudez, O.,, Livianos, L. Influence of stress in the onset of eating disorders: Data from a two-stage epidemiologic controlled study. Psychosom. Med. 2006, 68, 628–635. [CrossRef] [PubMed]

Russell CG, Russell A. Biological and Psychosocial Processes in the Development of Children’s Appetitive Traits: Insights from Developmental Theory and Research. Nutrients. 2018;10(6).

Schell, S. E., & Racine, S. E. (2023). Reconsidering the role of interpersonal stress in eating pathology: Sensitivity to rejection might be more important than actual experiences of peer stress. Appetite, 187, 106588

Sheyla C.S Fernandes, Ana Carolina Marinho, & Julio da Costa. (2023). ANALYSIS OF BELIEFS ABOUT BINGE EATING BEHAVIOR FROM THE THEORY OF PLANNED ACTION. New Trends in Qualitative Research, 18, e909. 10.36367/ntqr.18.2023.e909

Smith, K. E., Mason, T. B., Schaefer, L. M., Anderson, L. M., Critchley, K., Crosby, R. D., Engel, S. G., Crow, S. J., Wonderlich, S. A., & Peterson, C. B. (2021). Dynamic Stress Responses and Real-Time Symptoms in Binge-Eating Disorder. Annals of behavioral medicine : a publication of the Society of Behavioral Medicine, 55(8), 758–768. 10.1093/abm/kaaa061

Stice, E. (1998). Relations of restraint and negative affect to bulimic pathology: A longitudinal test of three competing models. International Journal of Eating Disorders, 23(3), 243–260.

Treasure, Janet; Duarte, Tiago Antunes; Schmidt, Ulrike (2020). Eating disorders. The Lancet, 395(10227), 899–911. doi:10.1016/S0140-6736(20)30059-3

Thornton LM, Trace SE, Brownley KA, Algars M, Mazzeo SE, Bergin JE, et al. A Comparison of Personality, Life Events, Comorbidity, and Health in Monozygotic Twins Discordant for Anorexia Nervosa. Twin Res Hum Genet. 2017;20(4):310–8. [PubMed: 28535840]

Walenda, A., Bogusz, K., Kopera, M., Jakubczyk, A., Wojnar, M., & Kucharska, K. (2021). Emotion regulation in binge eating disorder. Psychiatria Polska, 55(6), 1433–1448.

Wasserman, D.,, Iosue, M.,, Wuestefeld, A.,, Carli, V. Adaptation of evidence-based suicide prevention strategies during and after the COVID-19 pandemic. World Psychiatry 2020, 19, 294–306

